# MTS-Bench: A Manchester Triage System Benchmark for Language Model Triage Safety

**DOI:** 10.64898/2026.08.04.26359651

**Authors:** Sandhanakrishnan Ravichandran, Miguel Romano, Rogério Corga da Silva, Tiago Mendes, Nader Absi, Marta Isidoro, Shivesh Kumar, Michiel Van der Heijden, Valentine Emmanuel Gnanapragasam

## Abstract

**Background:** General purpose language models such as ChatGPT are increasingly used by physicians and triage nurses during emergency triage. A recent study reported 51.6% undertriage of emergencies when patients queried ChatGPT directly (Ramaswamy et al., 2026). DR. INFO is an agentic AI based clinical assistant that retrieves over a curated clinical knowledge base, and an MTS specific retrieval configuration is available in which the system also retrieves the Manchester Triage System (MTS) textbook at inference time. We evaluate all three configurations against the Manchester Triage System.

**Methods:** We adapted the clinical scenarios published by Ramaswamy et al. and mapped them to the Manchester Triage System, yielding 39 emergency cases covering all five MTS priority levels. Each case was evaluated in two variants, one without and one with the objective clinical data block (vital signs, examination findings, and laboratory results), and permuted across two genders, giving 156 prompts per system per condition (4 permutations of each of the 39 cases). Three systems were tested with and without a misleading GP referral statement prepended as an anchoring statement, giving 312 prompts per system: DR. INFO Baseline, DR. INFO with MTS retrieval, and OpenAI GPT-5.1. The primary outcome was the undertriage rate on the ordered MTS scale, tested with Fisher’s exact test.

**Results:** GPT-5.1 undertriaged 44.2% of prompts (69/156; 95% CI 36.7 to 52.1), including 75.0% of Red-priority and 73.4% of Orange-priority prompts. Both DR. INFO configurations undertriaged 11.5% of prompts (18/156; 95% CI 7.4 to 17.5; Fisher’s exact *p* = 1.0 *×* 10*^−^*^10^ versus GPT-5.1). The 156 prompts derive from 39 clinical cases (2 objective-data variants *×* 2 genders each); the 8 Red-priority prompts derive from 2 Red cases. When the anchoring statement was prepended, GPT-5.1 undertriaged 8 of 8 Red-priority prompts, while both DR. INFO configurations continued to undertriage none. Adding objective clinical data to the input reduced undertriage in DR. INFO with MTS retrieval from 19.2% to 3.8% (*p* = 0.005). DR. INFO Baseline and GPT-5.1 showed no comparable change. There was no statistically detectable gender difference.

**Conclusion:** On this benchmark, replacing a general purpose language model with an agentic retrieval augmented system over a curated clinical knowledge base substantially reduced the undertriage rate. Adding retrieval of the Manchester Triage System textbook to the agentic system was further associated with an appropriate change in the assigned MTS priority when objective clinical data became available. Of the three configurations evaluated here, DR. INFO with MTS retrieval combined a clinically conservative assignment at Red priority with a reduction in undertriage when objective clinical data were added (19.2% to 3.8%).

## 1 Introduction

Emergency triage stratifies patients by urgency under limited clinical resources. Errors are asymmetric: undertriage delays time critical care, whereas overtriage carries resource cost but rarely direct harm (Mackway-Jones et al., 2014). The initial acuity level is assigned at first contact, usually by a triage nurse and in some settings by a physician (small emergency departments (EDs), primary care, telephone triage), on partial information (chief complaint, brief examination, vital signs), and then revised as the picture evolves, vital signs change, and laboratory or imaging results return (Mackway-Jones et al., 2014). A Yellow MTS assignment can become an Orange or Red on reassessment if the trajectory worsens. The same pattern applies in primary care: a general practitioner (GP) referral letter often carries a tentative diagnosis that the receiving clinician must accept or revise, and the same case can look quite different on the letter and after a structured workup.

The Manchester Triage System (MTS) is a widely used nurse led emergency triage framework (Ingielewicz et al., 2025). It uses five priority levels with explicit time targets: Red (immediate, 0 min), Orange (very urgent, 10 min), Yellow (urgent, 60 min), Green (standard, 120 min), Blue (non urgent, 240 min) (Mackway-Jones et al., 2014). Each presentation is mapped to a chief complaint specific flowchart whose discriminators (e.g. “abnormal pulse”, “SpO_2_ below 95%”) are evaluated in fixed order from highest to lowest acuity; the first positive discriminator determines the priority. Inter rater agreement on vignette assessment is substantial (pooled reliability coefficient 0.751 (Mirhaghi et al., 2017); *κ* = 0.59 between front line and expert nurses, with approximately 28.6% of cases misclassified by experienced triage nurses (Zaboli et al., 2025)). The framework provides retrievable, time anchored ground truth: the correct MTS priority for an acute asthma exacerbation in respiratory distress is the same whether the case is first seen by a triage nurse, a primary care physician on a referral letter, or an ED physician after workup.

Large language models are increasingly used as a triage adjunct by clinicians. Performance on medical knowledge and dialogue based diagnostic tasks has improved rapidly (Singhal et al., 2023; Tu et al., 2025), and earlier evaluations characterised the triage performance of pre-GPT-4 and GPT-4 generation models on text vignettes (Levine et al., 2024; Kaboudi et al., 2024; Bedi et al., 2025). Uptake is now substantial: 66% of US physicians reported clinical use of AI tools in 2024 (AMA, 2024), and one in five UK general practitioners reports clinical use of an AI chatbot (Blease et al., 2024); qualitative work with ED physicians and nurses describes both growing acceptance and operational concerns about machine learning based triage (Güvey Emre et al., 2025). Two categories of tool are emerging in this space. General purpose language models such as ChatGPT are the most widely used; recent deployment evaluations have shown both encouraging results (an 18 percentage point improvement in physician clinical decision accuracy with GPT-4 assistance in a randomised trial (Goh et al., 2025); impact on real ED operations under prospective deployment (Taylor et al., 2025)) and reasons for caution (in a 39,375 patient retrospective, no current general purpose model reached *κ >* 0.80 with clinicians (Nedos et al., 2026)). The second category is the purpose built agentic AI based clinical assistant, which is not a general purpose chatbot but a system that retrieves over a curated clinical knowledge base and reasons over the retrieved evidence; a retrieval augmented triage workflow of this kind has been reported to reach 95.4% sensitivity for high risk patient detection (Wong et al., 2026). The most direct evidence of the safety gap between these categories comes from Ramaswamy et al. in *Nature Medicine* 2026, who reported 51.6% undertriage of true emergencies when patients queried ChatGPT directly, with anchoring statements increasing the probability of a triage shift more than tenfold (Ramaswamy et al., 2026). That study characterises the consumer self triage failure mode.

A concurrent independent comparison of frontier general purpose language models against two commercial clinical retrieval augmented tools on broad medical knowledge and real world physician query benchmarks reported that frontier models can match or exceed clinical retrieval augmented tools on those tasks (Vishwanath et al., 2026), framing the open question of whether and where structured retrieval contributes to clinical safety as a function of task type. The clinical question motivating this study is whether the same failure mode appears when a clinician uses these tools as a triage adjunct, against a recognised framework, on structured cases in the format clinicians actually receive. The objective of this study was to evaluate the safety of three AI triage adjuncts against the Manchester Triage System on the same set of structured clinical scenarios: a general purpose language model (OpenAI GPT-5.1, the same model family that powers the ChatGPT consumer product, accessed via API), an agentic retrieval augmented clinical assistant over a curated clinical knowledge base (DR. INFO Baseline), and the same agentic assistant with retrieval of the Manchester Triage System textbook added (DR. INFO with MTS retrieval). The clinical scenarios were adapted from Ramaswamy et al. and mapped to MTS priorities, then evaluated in two variants (without and with the objective clinical data block of vital signs, examination findings, and laboratory results) and permuted across two genders. Each case was also evaluated with and without a misleading GP referral statement prepended, to test resistance to authoritative but incorrect framing. The benchmark evaluates triage adjuncts, not replacements: in person triage involves direct physiological observation and experience informed judgement that text based systems cannot replicate, and the role envisaged for either category here is to support, not make, the clinician’s triage decision.

## 2 Methods

### 2.1 MTS-Bench dataset

MTS-Bench comprises 39 emergency vignettes adapted from the clinical scenario dataset of Ramaswamy et al. (drawn from the primary set of 30 scenarios and the expanded supplementary set of 9 additional scenarios covering textbook emergencies and additional psychiatric presentations) (Ramaswamy et al., 2026). The 39 cases span 19 clinical domains after normalisation of near-synonymous domain labels in the source dataset. Each case is provided in the structured clinical format physicians and triage nurses receive in practice (chief complaint, history of present illness, relevant past medical history, and, where applicable, objective data) at two variants: Version F (without objective data, i.e. history only) and Version E (with objective data, defined per Ramaswamy et al. as laboratory values, vital signs, and physical examination findings). The two versions share identical clinical text apart from the objective data block, isolating the effect of objective clinical information on triage assignment within each scenario. Each variant was permuted across two genders to give 156 prompts per anchoring condition (Figure 1). To test susceptibility to clinical anchoring, each vignette was also paired with a misleading GP referral statement (“Referred by GP who suspects [benign diagnosis]”) prepended to the presentation. The benign diagnosis for each case was authored by a clinician to be plausible but incorrect, such that uncritical acceptance of the referral framing would produce undertriage. This design adapts the anchoring methodology of Ramaswamy et al. (Ramaswamy et al., 2026): their family member minimisation framing is replaced with a professional clinical framing that reflects the form clinicians encounter in routine practice (Croskerry, 2013). The full anchor list is available from the authors on request. The case mix spans all five MTS priority levels and includes presentations with both classical and trajectory dependent acuity (Table 1). Each case was mapped to an MTS priority through a two stage process for fidelity to the Manchester Triage Group reference text (Mackway-Jones et al., 2014): an assignment pipeline identified the appropriate presentational flowchart and evaluated the discriminators from Red through Blue, assigning the priority corresponding to the first positive discriminator, and a verification pipeline cross referenced each assignment against the textbook with explicit page references and discriminator quotations. Every assignment was then reviewed and verified by the physician co-authors against the same reference text. All 39 final priorities reflect physician verified consensus. The DR. INFO with MTS retrieval configuration retrieves passages from the same Manchester Triage Group reference text used by the annotation pipeline (see Section 5). The study used these clinician authored vignettes only and did not involve human subjects, real clinical data, or patient interaction, and did not require ethics approval.

**Figure 1:**
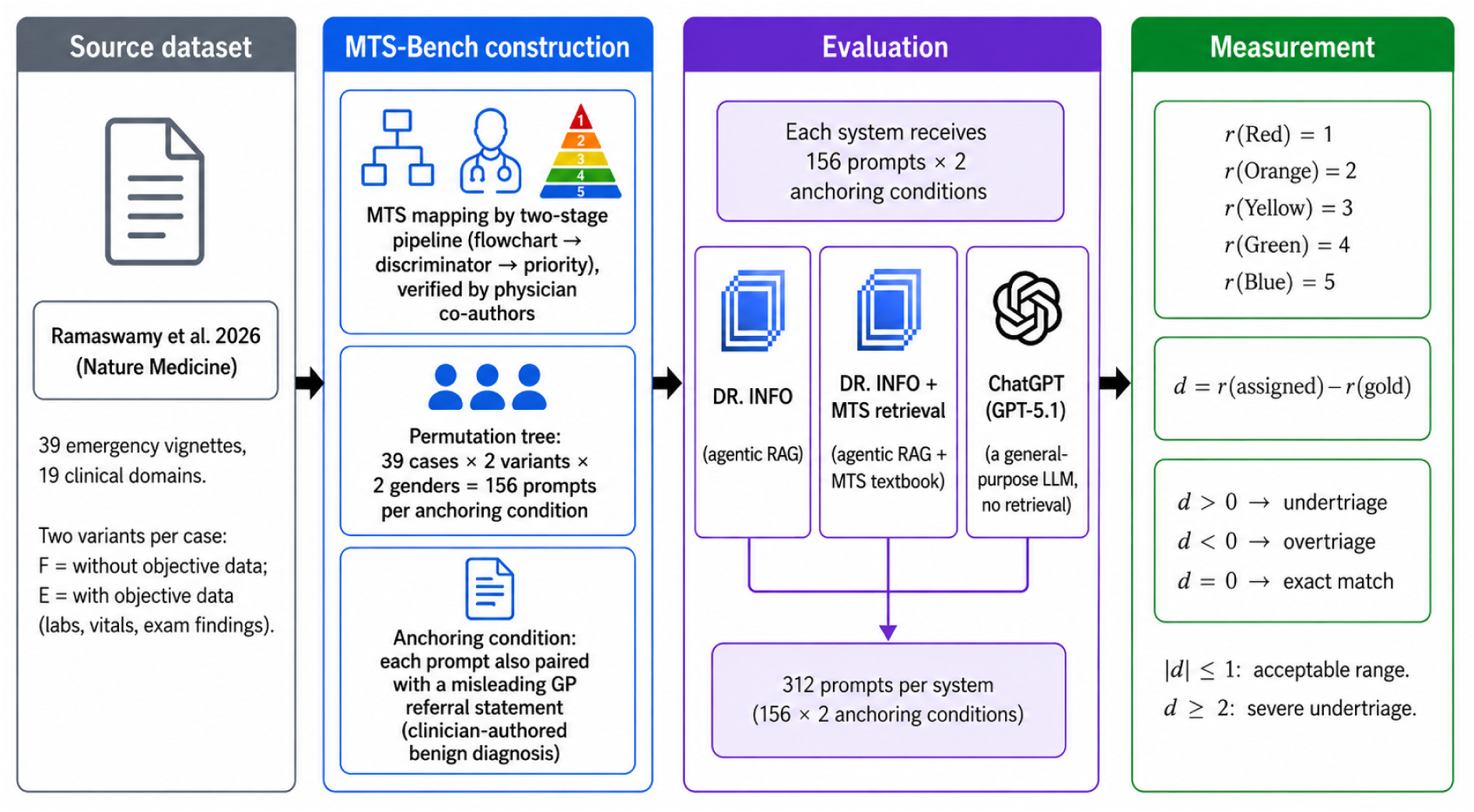
Study design and dataset construction. The 39 vignette set (spanning 19 clinical domains) was adapted from the Ramaswamy et al. 2026 clinical scenario dataset (30 primary scenarios and 9 additional scenarios from the expanded supplementary set) and mapped to the MTS five level priority scheme, permuted across two case variants (with and without objective data) and two genders to give 156 prompts per anchoring condition, and evaluated under two anchoring conditions (with and without a misleading GP referral statement). Three systems received 312 prompts each. Outcomes were derived from a rank function *r* on the MTS priorities and the difference *d* = *r*(assigned) *− r*(gold).

**Table 1:** Distribution of MTS-Bench cases by gold-standard MTS priority. Permutations per priority is the number of evaluated prompts per system per anchoring condition (cases *×* 2 case variants *×* 2 genders).

| MTS priority | Target time | Cases ($n$) | Permutations ($n$) |
| --- | --- | --- | --- |
| ■ Red (Immediate) | 0 min | 2 | 8 |
| ■ Orange (Very urgent) | 10 min | 16 | 64 |
| ■ Yellow (Urgent) | 60 min | 13 | 52 |
| ■ Green (Standard) | 120 min | 6 | 24 |
| ■ Blue (Non urgent) | 240 min | 2 | 8 |
| **Total** |  | **39** | **156** |

### 2.2 Systems evaluated

Three systems were evaluated on the same 312 prompts per system (156 per anchoring condition). DR. INFO is an agentic AI based clinical assistant: for a given clinical query it decomposes the question, retrieves evidence iteratively from a curated knowledge base of medical literature and guidelines, and produces a structured clinical assessment with citations. The curated knowledge base used in the Baseline configuration consists of general clinical literature and practice guidelines and does not include the Manchester Triage Group reference text used to construct the gold standard for this benchmark. Prior evidence on DR. INFO comprises a prospective point of care pilot in which physicians evaluated the system on real clinical queries (Corga Da Silva et al., 2026), and a benchmark evaluation on the OpenAI HealthBench realistic clinical query set (Ravichandran et al., 2025). The MTS retrieval configuration adds a retrieval layer dedicated to the Manchester Triage System reference text (Mackway-Jones et al., 2014), returning the relevant presentational flowcharts, discriminator definitions, and decision logic for each query. OpenAI GPT-5.1 was accessed via the OpenAI application programming interface (API), with a standardised system prompt instructing the model to perform MTS-based triage assessment. GPT-5.1 is the same model family that powers the ChatGPT consumer product; we evaluated the model via API rather than through the ChatGPT interface to allow controlled prompting, consistent parameters across runs, and higher throughput than the consumer inter-face permits. Results reported here therefore apply to GPT-5.1 as accessed via API and may differ from the ChatGPT consumer product behaviour, which includes additional server-side guardrails and prompt scaffolding not exposed via the API. The DR. INFO Baseline versus DR. INFO with MTS retrieval contrast isolates the effect of MTS specific retrieval within the same agentic system. The DR. INFO versus GPT-5.1 contrast reflects the comparison between two classes of language model based clinical tool that clinicians have access to.

### 2.3 Evaluation, outcomes, and statistics

Each prompt was submitted once per system per condition via API. Triage assignments were compared against the gold standard on the ordered MTS scale, which we index from highest to lowest acuity as

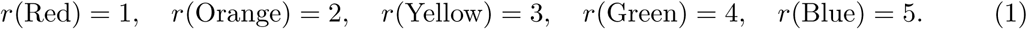

For a response with gold priority *g* and assigned priority *a*, we define

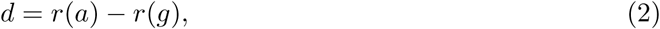

and classify the outcome as undertriage if *d >* 0, overtriage if *d <* 0, and exact match if *d* = 0. Within one level agreement required *|d| ≤* 1; severe undertriage required *d ≥* 2. Rates were stratified by priority, case variant, anchoring condition, and gender.

Each rate (undertriage, overtriage, and so on) is reported with a Wilson 95% confidence interval (Wilson, 1927). Differences between systems were tested with Fisher’s exact test (two sided) as the primary analysis. The 156 permutations per system are 4 permutations (2 case variants *×* 2 genders) of 39 underlying clinical cases and are therefore not independent observations. To check that the between-system contrasts do not depend on this assumption, we ran two case-level sensitivity analyses: (i) a case-level Fisher’s exact test in which each case (n=39) was coded as undertriaged if a majority of its 4 permutations were undertriaged, and (ii) a generalised estimating equations logistic regression on the full 156 permutations with an exchangeable working correlation clustered by case. Reporting follows the STARD recommendations (Cohen et al., 2016).

### 2.4 Independent physician adjudication

#### Purpose

To provide an external check on the gold standard assignments produced by the inhouse pipeline, the high risk subset of prompts generated by the three systems was additionally re-evaluated by an independent physician.

#### Inclusion criteria

A prompt was included in the adjudication subset if it met either of two automated high risk criteria across all three systems and both anchoring conditions: Red or Orange gold standard undertriage, or severe undertriage of *≥* 2 MTS levels.

#### Sample composition

Applying these criteria to the full evaluation set of 936 system responses (three systems *×* 156 prompts *×* two anchoring conditions) yielded 151 prompts for adjudication, distributed across systems in proportion to how often each system met the high risk criteria: 24 prompts from DR. INFO Baseline, 20 from DR. INFO with MTS retrieval, and 107 from GPT-5.1. The per-system counts therefore differ substantially. This imbalance is a direct consequence of the high risk subset design: each system contributed prompts in proportion to how often it met the automated high risk criteria, and the relative frequency at which each system was flagged is reported as an upstream finding in its own right in Section 3.3.

#### Blind review procedure

The 151 prompts were presented to the reviewer through a dedicated web interface in a single blinded protocol. The reviewer was a physician unaffiliated with the development of DR. INFO. For each prompt, the reviewer was shown the full text sent to the system and the full clinical reasoning text returned by the system, and was asked to assign the MTS priority that he would have given the presentation in clinical practice. The reviewer was blinded to two pieces of information: the identity of the system that had produced each response (DR. INFO Baseline, DR. INFO with MTS retrieval, or GPT-5.1), and the MTS-Bench gold standard priority assigned by the in-house pipeline. The 151 prompts were presented in randomised order with a stratification constraint that prevented consecutive presentation of two permutations of the same underlying clinical scenario, so that system identity could not be inferred from clustering. Reviewer responses were saved locally on every change and exported as a single comma separated file at the end of the review.

#### Analysis

Agreement between the physician and the system assigned priority was summarised by raw agreement and by quadratic weighted Cohen’s kappa (Cohen, 1968), the latter being the standard agreement metric for ordinal scales such as MTS. The direction of any disagreement was classified as a physician confirmed under call (the physician assigned a higher acuity than the system, the clinically dangerous direction) or a physician confirmed over call (the physician assigned a lower acuity than the system, the clinically safer direction). Both the upstream rate at which each system met the high risk criteria and the within-subset physician confirmed under call rate are reported with Wilson 95% confidence intervals (Wilson, 1927). Results are reported in Section 3.3.

## 3 Results

Each system was evaluated on 312 prompts (156 prompts in each of two conditions: with and without a misleading GP referral statement prepended as an anchoring statement). All systems returned a parseable MTS priority for every prompt, with no failures.

### 3.1 Anchoring bias

We first describe performance without the anchoring statement, then contrast it with performance when each prompt was prepended with a misleading one-line GP referral of the form “Referred by GP who suspects [benign diagnosis]”.

#### Without anchoring statement

Both DR. INFO configurations undertriaged 11.5% of prompts (18/156); GPT-5.1 undertriaged 44.2% (69/156). Exact-match accuracy was 40.4% for DR. INFO Baseline, 44.9% for DR. INFO with MTS retrieval, and 50.6% for GPT-5.1. The error directions differed: DR. INFO errors were predominantly overtriage (48.1% Baseline, 43.6% MTS retrieval) while GPT-5.1 errors were predominantly undertriage with only 5.1% overtriage. Full performance and between-system test statistics are given in Table 2. The case-level sensitivity analyses pre-specified in Section 2.3 were consistent with the permutation level Fisher’s exact test: in the case-level analysis, 3 of 39 cases were majority undertriaged by DR. INFO Baseline, 1 of 39 by DR. INFO with MTS retrieval, and 16 of 39 by GPT-5.1 (case-level Fisher’s exact GPT-5.1 vs DR. INFO Baseline *p* = 0.001; GPT-5.1 vs DR. INFO with MTS retrieval *p <* 0.001). In the GEE logistic regression clustered by case on the full 156 permutations, GPT-5.1 remained significantly more likely to undertriage than each DR. INFO configuration (*p <* 10*^−^*^6^ in both contrasts).

**Table 2:**
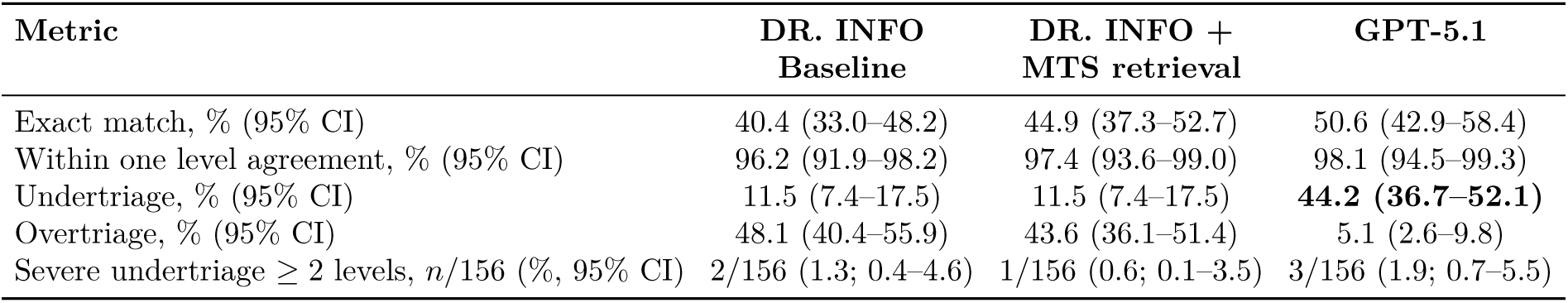
Triage performance without anchoring statement, *n* = 156 per system. Metrics use the index-based definitions in Section 2.3. Within one level agreement is *±*1 MTS level of the gold standard. Bold marks the worst value in each safety-relevant row. Between-system test (Fisher’s exact, two-sided): undertriage of GPT-5.1 vs DR. INFO Baseline, 44.2% vs 11.5%, OR 6.08 (95% CI 3.39–10.90; DR. INFO Baseline as reference), *p* = 1.0 *×* 10*^−^*^10^.

| Metric | DR. INFO<br>Baseline | DR. INFO +<br>MTS retrieval | GPT-5.1 |
| --- | --- | --- | --- |
| Exact match, % (95% CI) | 40.4 (33.0–48.2) | 44.9 (37.3–52.7) | 50.6 (42.9–58.4) |
| Within one level agreement, % (95% CI) | 96.2 (91.9–98.2) | 97.4 (93.6–99.0) | 98.1 (94.5–99.3) |
| Undertriage, % (95% CI) | 11.5 (7.4–17.5) | 11.5 (7.4–17.5) | **44.2 (36.7–52.1)** |
| Overtriage, % (95% CI) | 48.1 (40.4–55.9) | 43.6 (36.1–51.4) | 5.1 (2.6–9.8) |
| Severe undertriage $\geq 2$ levels, $n/156$ (%, 95% CI) | 2/156 (1.3; 0.4–4.6) | 1/156 (0.6; 0.1–3.5) | 3/156 (1.9; 0.7–5.5) |

Stratification by gold-standard MTS priority is shown in Figure 2 and Table 3. At the two highest acuity levels (Red and Orange), GPT-5.1 undertriaged the majority of prompts (75.0% at Red and 73.4% at Orange) while both DR. INFO configurations undertriaged none at Red and about 20% at Orange. The 8 Red-priority prompts and the 64 Orange-priority prompts derive from 2 Red cases and 16 Orange cases respectively. At the lower acuity levels (Yellow, Green, Blue), undertriage was rare across all three systems, with the exception of Green for GPT-5.1 (50.0%). Classified by severity (Table 4), undertriage events in all three systems were dominated by Orange-priority prompts being downgraded to a lower priority; in GPT-5.1 this category alone accounted for 47 of the 69 total undertriage events. At the Red priority, the 6 GPT-5.1 undertriage events without the anchoring statement split 4 + 2 across the two Red cases (aortic dissection, undertriaged in 4 of 4 permutations; anaphylaxis with airway compromise, undertriaged in 2 of 4 permutations, both from the variant with objective data). All 32 Red-priority evaluations (2 systems *×* 2 conditions *×* 2 cases *×* 4 permutations) of DR. INFO Baseline and DR. INFO with MTS retrieval were correctly triaged as Red.

**Figure 2:**
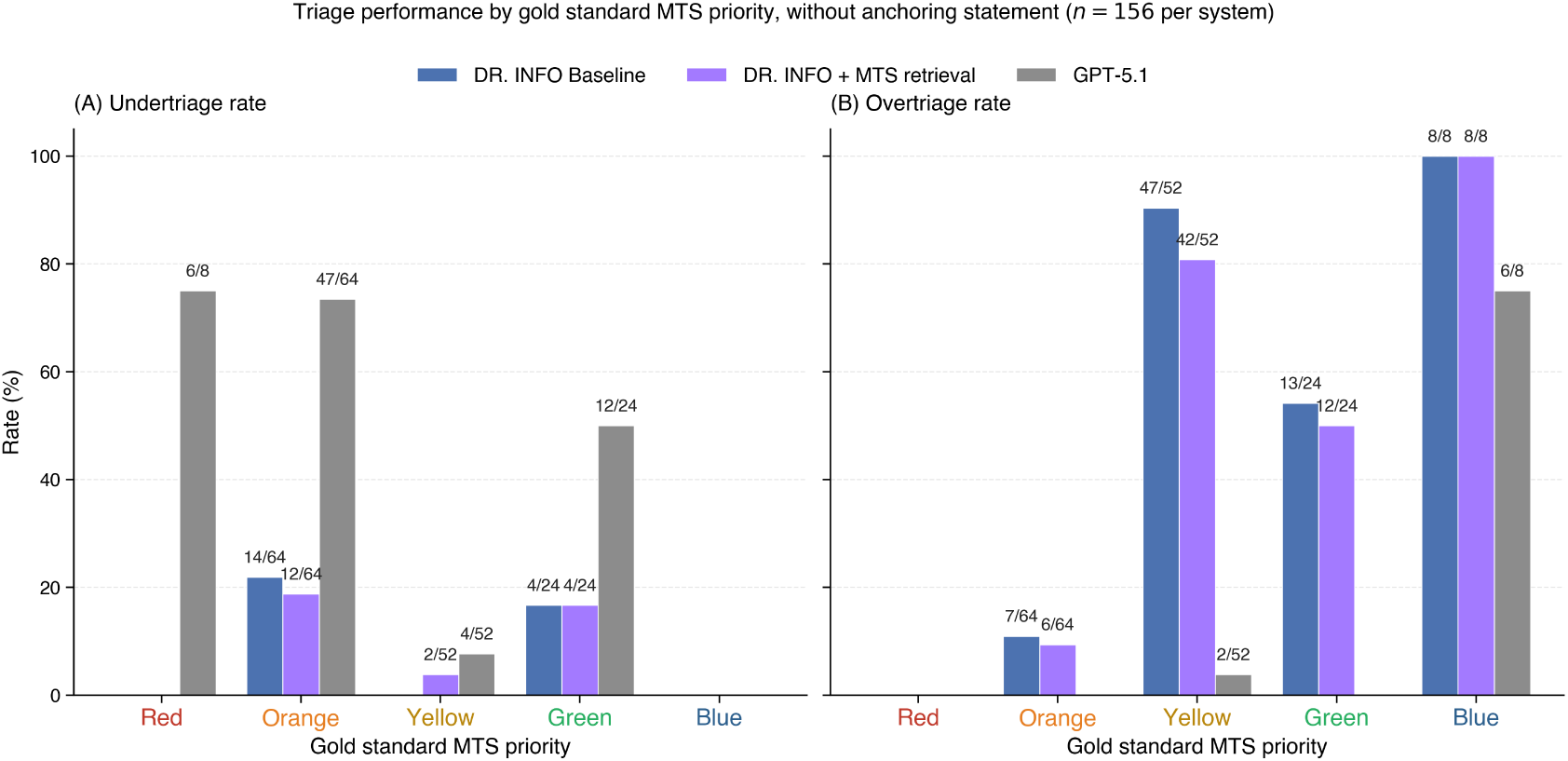
Triage performance by gold-standard MTS priority, without anchoring statement (*n* = 156 per system). (A) Undertriage rate. (B) Overtriage rate. Numerator/denominator counts are annotated above each bar.

**Table 3:** Undertriage rate by gold-standard MTS priority, without anchoring statement, with Wilson 95% confidence intervals. Denominators are the number of prompts at each priority (Red=8 prompts from 2 cases; Orange=64 from 16; Yellow=52 from 13; Green=24 from 6; Blue=8 from 2; 4 permutations per case). Bold marks the worst value per row.

| Gold standard priority | DR. INFO Baseline<br>% (95% CI), $n/N$ | DR. INFO + MTS retrieval<br>% (95% CI), $n/N$ | GPT-5.1<br>% (95% CI), $n/N$ |
| --- | --- | --- | --- |
| ■ **Red (Immediate)** | 0.0 (0.0–32.4), 0/8 | 0.0 (0.0–32.4), 0/8 | **75.0 (40.9–92.9), 6/8** |
| ■ **Orange (Very urgent)** | 21.9 (13.5–33.4), 14/64 | 18.8 (11.1–30.0), 12/64 | **73.4 (61.5–82.7), 47/64** |
| ■ **Yellow (Urgent)** | 0.0 (0.0–6.9), 0/52 | 3.8 (1.1–13.0), 2/52 | 7.7 (3.0–18.2), 4/52 |
| ■ **Green (Standard)** | 16.7 (6.7–35.9), 4/24 | 16.7 (6.7–35.9), 4/24 | 50.0 (31.4–68.6), 12/24 |
| ■ **Blue (Non urgent)** | 0.0 (0.0–32.4), 0/8 | 0.0 (0.0–32.4), 0/8 | 0.0 (0.0–32.4), 0/8 |

**Table 4:**
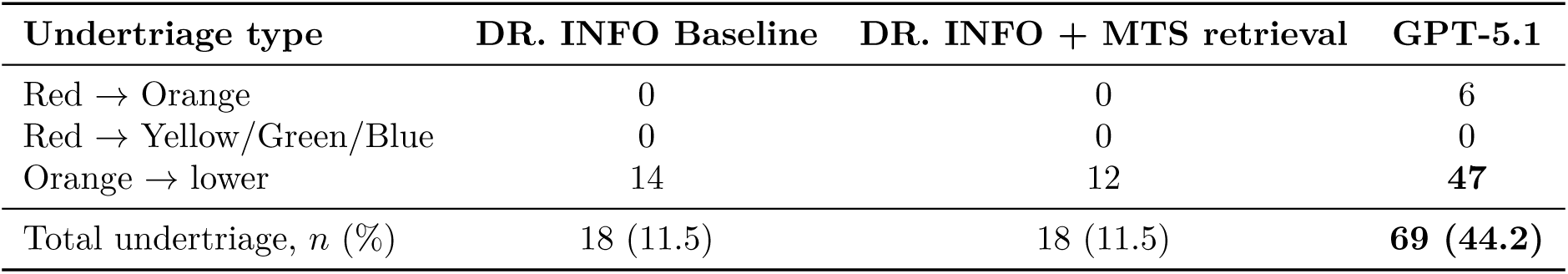
Undertriage events classified by clinical severity, without anchoring statement, *n* = 156 prompts per system. The three severity rows include only Red-origin and Orange-origin events. Yellow-origin and Green-origin undertriage (16 events for GPT-5.1, 4 for DR. INFO Baseline, 6 for DR. INFO with MTS retrieval) are not classified here. Bold marks the worst value per row.

| Undertriage type | DR. INFO Baseline | DR. INFO + MTS retrieval | GPT-5.1 |
| --- | --- | --- | --- |
| Red → Orange | 0 | 0 | 6 |
| Red → Yellow/Green/Blue | 0 | 0 | 0 |
| Orange → lower | 14 | 12 | **47** |
| Total undertriage, $n$ (%) | 18 (11.5) | 18 (11.5) | **69 (44.2)** |

#### With anchoring statement

The overall undertriage rate did not change significantly for any system (Table 5). At the Red priority, GPT-5.1 undertriaged 8/8 prompts with the anchoring statement compared with 6/8 without it (Table 5). The two DR. INFO configurations remained at 0/8 Red-priority prompts undertriaged. The 8 Red-priority prompts per condition derive from 2 Red cases (4 permutations each); with the anchoring statement, GPT-5.1 undertriaged all 4 permutations of both Red cases. Figure 3 shows the priority-stratified pattern under the anchoring statement.

**Figure 3:**
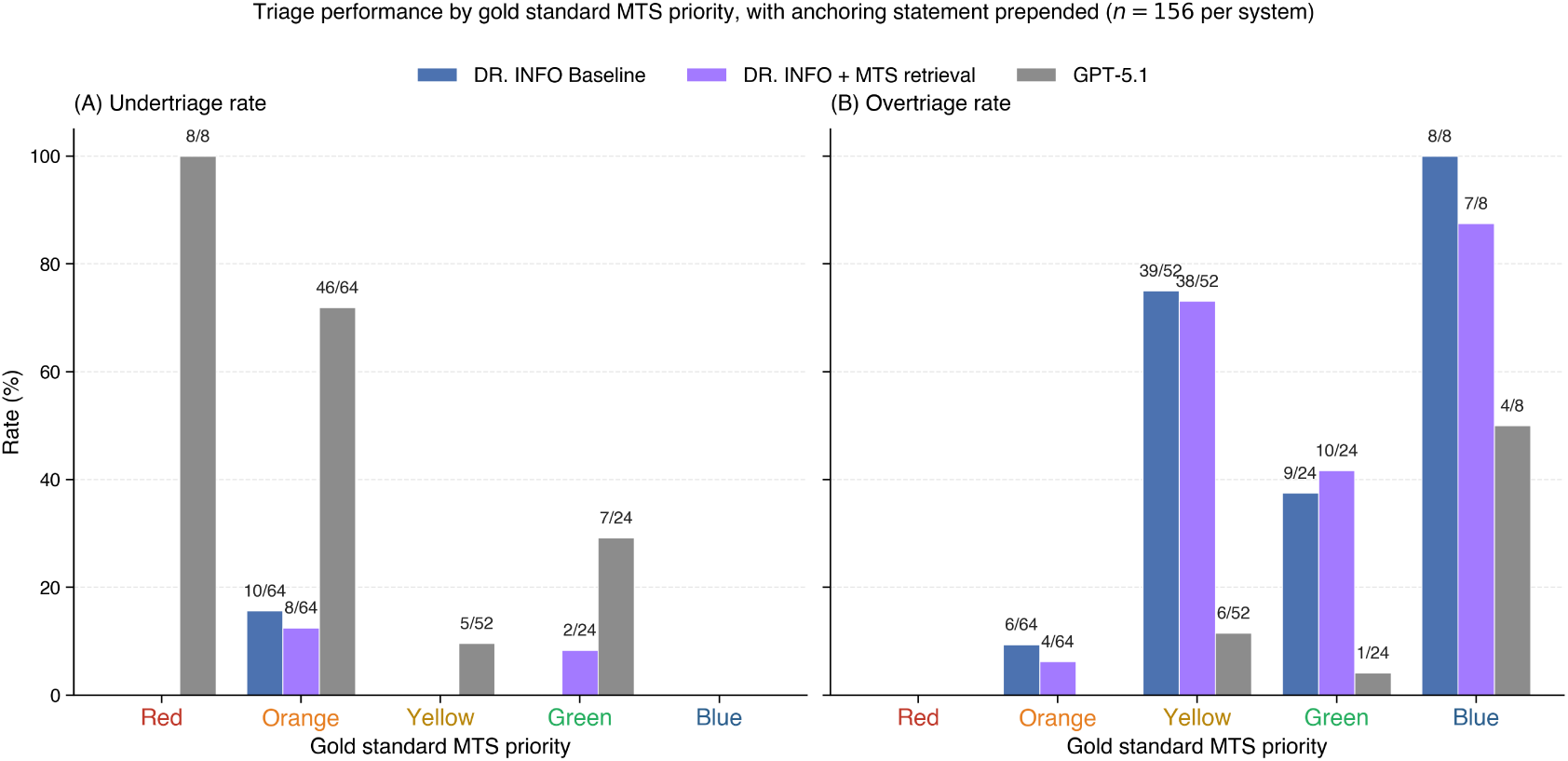
Triage performance by gold-standard MTS priority, with the misleading GP referral statement prepended as the anchoring statement (*n* = 156 per system). (A) Undertriage rate. (B) Overtriage rate. Numerator/denominator counts are annotated above each bar.

**Table 5:** Triage performance with and without the misleading GP referral statement, *n* = 156 per condition per system. Within-system *p* values are from Fisher’s exact test (two-sided). Bold marks the worst value per row.

|  | Condition | DR. INFO Baseline | DR. INFO + MTS retrieval | GPT-5.1 |
| --- | --- | --- | --- | --- |
| Undertriage, $n/156$ (%) | No referral | 18/156 (11.5) | 18/156 (11.5) | 69/156 (44.2) |
|  | With referral | 10/156 (6.4) | 10/156 (6.4) | 66/156 (42.3) |
| | $p$ value | 0.165 | 0.165 | 0.819 |
| Red undertriage, $n/8$ (%) | No referral | 0/8 (0.0) | 0/8 (0.0) | 6/8 (75.0) |
|  | With referral | 0/8 (0.0) | 0/8 (0.0) | **8/8 (100.0)** |
| Red → Orange, $n$ | No referral | 0 | 0 | 6 |
|  | With referral | 0 | 0 | **8** |
| Orange → lower, $n$ | No referral | 14 | 12 | 47 |
|  | With referral | 10 | 8 | 46 |

There was no statistically detectable gender difference in undertriage in any system, under either condition. Without the anchoring statement, undertriage in men versus women was 10.3% vs 12.8% (DR. INFO Baseline, *p* = 0.803), 11.5% vs 11.5% (DR. INFO with MTS retrieval, *p* = 1.000), and 47.4% vs 41.0% (GPT-5.1, *p* = 0.519). With the anchoring statement, undertriage in men versus women was 7.7% vs 5.1% (DR. INFO Baseline, *p* = 0.746), 9.0% vs 3.8% (DR. INFO with MTS retrieval, *p* = 0.327), and 41.0% vs 43.6% (GPT-5.1, *p* = 0.871). All *p* values are from Fisher’s exact test (two-sided), with *n* = 78 per gender per condition.

### 3.2 Additional clinical context: vital signs, examination findings, laboratory results

Each case was evaluated in two variants (Section 2.1): without objective clinical data (Version F) and with objective clinical data added (Version E).

For DR. INFO + MTS retrieval, undertriage was 19.2% (15/78) on the variant without objective data and 3.8% (3/78) on the variant with objective data (Table 6, Figure 4). The reduction was distributed across Orange (9/32 → 3/32), Yellow (2/26 → 0/26), and Green (4/12 → 0/12). DR. INFO Baseline changed in the same direction with a smaller, non significant difference (12.8% → 10.3%). GPT-5.1 changed in the opposite direction, with undertriage rising from 41.0% (32/78) on the variant without objective data to 47.4% (37/78) on the variant with objective data. When the same comparison was repeated with the anchoring statement prepended, the reduction in undertriage between the variant without objective data and the variant with objective data was smaller in all three systems (Figure 5).

**Figure 4:**
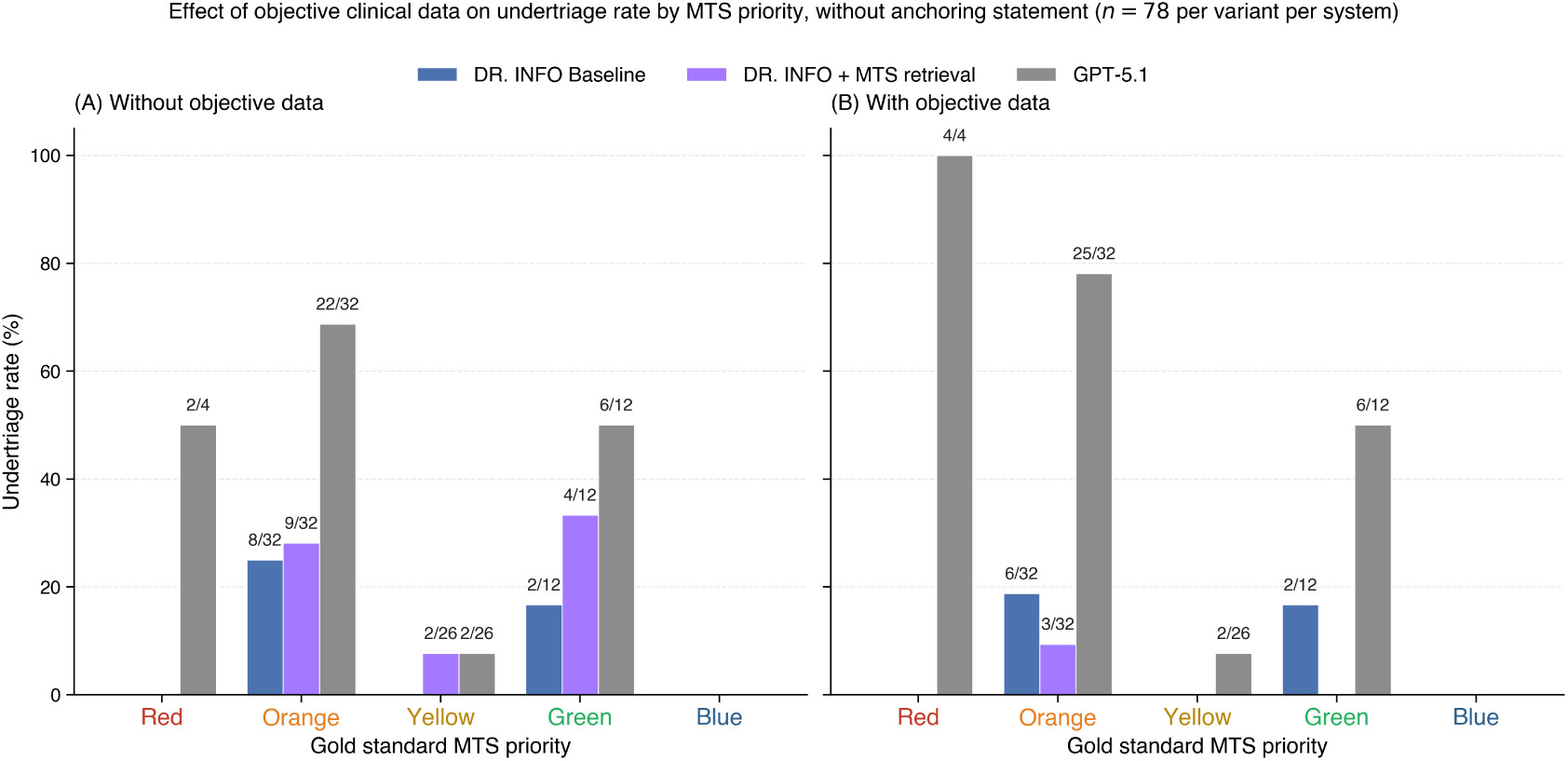
Effect of adding objective clinical data on undertriage rate by gold-standard MTS priority, without anchoring statement (*n* = 78 per variant per system). (A) Undertriage rate on the variant without objective data. (B) Undertriage rate on the same cases with the objective data block added. Numerator/denominator counts are annotated above each bar.

**Figure 5:**
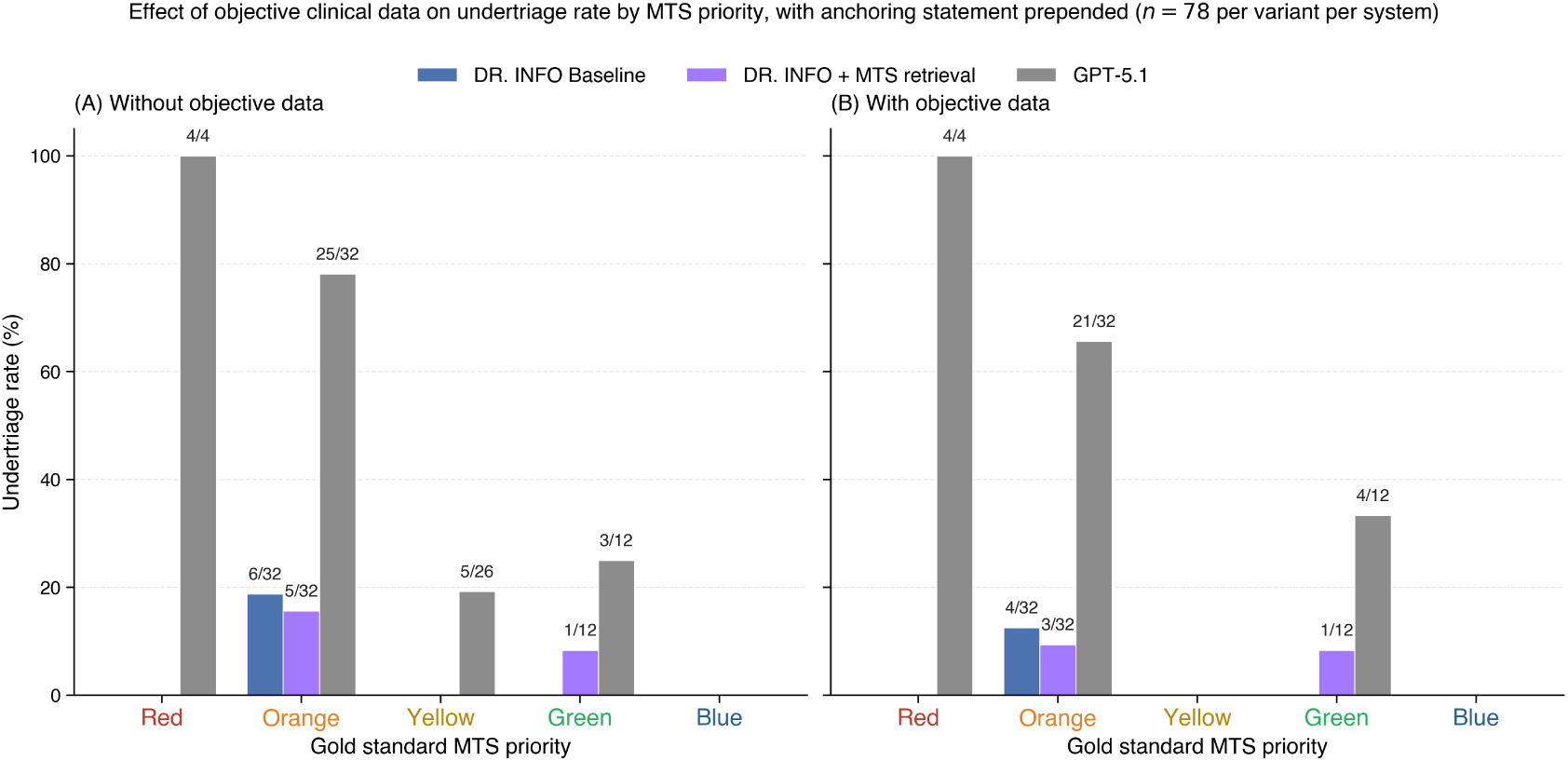
Effect of adding objective clinical data on undertriage rate by gold-standard MTS priority, with anchoring statement prepended (*n* = 78 per variant per system). (A) Undertriage rate on the variant without objective data. (B) Undertriage rate on the same cases with the objective data block added. The within-system reduction between (A) and (B) is smaller in all three systems than under the no-anchoring condition shown in Figure 4. Numerator/denominator counts are annotated above each bar.

**Table 6:** Effect of adding objective clinical data (vital signs, examination findings, and laboratory results) on triage performance, without anchoring statement. *n* = 78 per variant per system. Within-system *p* values from Fisher’s exact test (two-sided). For DR. INFO + MTS retrieval, OR 0.17 (with objective data versus without; variant without objective data as reference). Bold marks statistically significant differences within a system.

| Metric | Variant | DR. INFO Baseline | DR. INFO + MTS retrieval | GPT-5.1 |
| --- | --- | --- | --- | --- |
| Undertriage, % (95% CI) | With objective data (E) | 10.3 (5.1–19.4) | **3.8 (1.3–10.7)** | 47.4 (36.7–58.4) |
|  | Without (F) | 12.8 (7.1–21.8) | 19.2 (12.0–29.3) | 41.0 (30.6–52.4) |
|  | *p* value | 0.803 | **0.005** | 0.519 |

### 3.3 Independent physician adjudication of high-risk prompts

The physician adjudication produced two separable findings.

First, the rate at which each system met the automated high risk criteria differed substantially across systems (Table 7). Of the 312 responses generated per system across both anchoring conditions, GPT-5.1 met the high risk criteria in 107 prompts (34.3%; Wilson 95% CI 29.2 to 39.7), compared with 24 prompts for DR. INFO Baseline (7.7%; 5.2 to 11.2) and 20 prompts for DR. INFO with MTS retrieval (6.4%; 4.2 to 9.7). The confidence intervals do not overlap, indicating that the upstream automated pipeline flagged GPT-5.1 responses as high risk at approximately four to five times the rate of either DR. INFO configuration.

**Table 7:** Independent physician adjudication. Left column: how often each system met the automated high risk criteria (Red or Orange undertriage, or severe undertriage of *≥* 2 MTS levels) per 312 prompts. Right column: within the high risk subset, the proportion of prompts for which the physician assigned a higher acuity than the system (a physician confirmed under call, the clinically dangerous direction). Confidence intervals are computed by the Wilson method. In both columns the GPT-5.1 confidence interval does not overlap with the DR. INFO (combined) confidence interval.

| System | High risk prompts / 312 responses<br>(%, Wilson 95% CI) | Physician confirmed under calls<br>within the high risk subset (%, Wilson 95% CI) |
| --- | --- | --- |
| DR. INFO Baseline | 24/312 (7.7; 5.2–11.2) | 0/24 (0.0; 0.0–13.8) |
| DR. INFO with MTS retrieval | 20/312 (6.4; 4.2–9.7) | 0/20 (0.0; 0.0–16.1) |
| DR. INFO (combined) | 44/624 (7.1; 5.3–9.4) | 0/44 (0.0; 0.0–8.0) |
| GPT-5.1 | **107/312 (34.3; 29.2–39.7)** | **15/107 (14.0; 8.7–21.8)** |

Second, within the high risk subset the physician confirmed an under call (the physician assigned a higher acuity than the system, the clinically dangerous direction) in 15 of 107 GPT-5.1 prompts (14.0%; 95% CI 8.7 to 21.8) and in 0 of 44 DR. INFO prompts across both configurations combined (0.0%; 95% CI 0.0 to 8.0). The confidence intervals do not overlap. The physician adjudication therefore independently confirmed that the upstream pipeline was not over-flagging GPT-5.1 prompts, and that the safety asymmetry between the standalone language model and the retrieval augmented systems is present at both the automated and the physician adjudicated levels of evidence.

Two further descriptive statistics are reported for completeness. Per-system raw agreement between the physician and the system assigned priority was 90.0% (18 of 20 prompts) for DR. INFO with MTS retrieval, 83.3% (20 of 24) for DR. INFO Baseline, and 84.1% (90 of 107) for GPT-5.1; the corresponding quadratic weighted Cohen’s kappa was 0.46, 0.57, and 0.71 respectively, with the GPT-5.1 figure comparable to the kappa of 0.67 previously reported for GPT-4 against MTS consensus on a separate 124-case vignette set (Zaboli et al., 2024). The raw agreement and the weighted kappa order the systems differently: raw agreement is highest for DR. INFO with MTS retrieval, and weighted kappa is highest for GPT-5.1. The two statistics answer different questions on this subset. Raw agreement counts the proportion of prompts on which the physician and the system assigned the same priority. Weighted kappa also rewards small disagreements less harshly than large disagreements and penalises agreement expected under the observed marginal distributions. In a small high risk subset with skewed marginals (20 prompts for DR. INFO with MTS retrieval, most concentrated at one or two priority levels), the expected agreement term is unstable, and weighted kappa should be interpreted as descriptive rather than as a stable point estimate of ordinal agreement. The physician adjudicated over call rate (the physician assigned a lower acuity than the system, the clinically safer direction) was 4 of 24 (16.7%) for DR. INFO Baseline, 2 of 20 (10.0%) for DR. INFO with MTS retrieval, and 2 of 107 (1.9%) for GPT-5.1.

A short clinical commentary from the reviewing physician summarised the pattern as follows: the DR. INFO disagreements were generally toward greater caution (over call) rather than toward false reassurance, whereas the GPT-5.1 disagreements were toward missing presentations for which a lower priority assignment would be clinically uncomfortable. The clinical scenarios that the reviewer flagged most strongly were suspected diabetic ketoacidosis in type 1 diabetes, severe hypertension without focal neurological symptoms, and suicidal ideation with a named method; in each of these the reviewer would have assigned a higher priority than GPT-5.1 did, on the basis that delayed clinical review can change the safety outcome. The independent adjudication therefore confirmed the safety asymmetry observed in the automated analysis, with raw physician agreement highest for DR. INFO with MTS retrieval on this subset (with the weighted kappa caveat noted above).

## 4 Discussion

The three systems evaluated in this benchmark correspond to three increasingly grounded reasoning regimes for a triage adjunct: a general purpose language model that reasons from its own parameters alone (OpenAI GPT-5.1, the same model family that powers the ChatGPT consumer product, accessed via API), an agentic retrieval-augmented system that reasons over a curated clinical knowledge base (DR. INFO Baseline), and the same agentic system retrieving the Manchester Triage System textbook itself (DR. INFO with MTS retrieval). The data show the contribution of each step to triage safety.

### 4.1 Safety profile of the general purpose language model

GPT-5.1’s errors on this benchmark were not distributed symmetrically around the gold standard but were concentrated at the Red (Immediate) and Orange (Very urgent) priorities and were directional toward undertriage, meaning that the model assigned a lower MTS priority than the gold standard on the highest-acuity vignettes. The anchoring contrast further showed that a brief, clinician authored framing prepended to the case did not leave the safety profile unchanged but instead amplified undertriage at the Red priority, the level at which the time target is most demanding (0 minutes; immediate assessment). The qualitative pattern is similar in direction to the consumer self-triage failure reported by Ramaswamy et al. for ChatGPT on a different acuity scale (Ramaswamy et al., 2026), and is consistent with a multicentre retrospective in which no current general purpose model reached *κ >* 0.80 with clinician triage in 39,375 patients (Nedos et al., 2026). Because our scenarios were adapted from the Ramaswamy case set, the agreement is informative about how the failure mode survives a change of acuity scale and a change of anchoring framing, rather than being independent replication. One plausible mechanism is central tendency bias: a model trained on a corpus that overrepresents intermediate-acuity clinical advice and underrepresents structured, time-anchored emergency decision-making would be expected to regress its predictions toward the modal acuity of the training distribution, producing undertriage at the high-acuity tail and overtriage at the low-acuity tail. The inverted U-shaped accuracy pattern Ramaswamy et al. reported for ChatGPT is the same shape. Whether central tendency fully explains the pattern, or whether other mechanisms also contribute (instruction tuning toward reassurance, consumer-facing safety guardrails biased toward lower-acuity outputs), requires further investigation. The practical implication is the same in either case: closing the safety gap appears to require structured decision logic at inference time, not at training time.

### 4.2 Effect of agentic retrieval over a curated clinical knowledge base

An agentic retrieval augmented system reasoning over a curated clinical knowledge base substantially reduced the safety relevant errors observed in GPT-5.1, even without any framework specific tuning. Both DR. INFO configurations produced the same overall undertriage rate without the anchoring statement, and no Red undertriage under any condition, including with the misleading GP referral statement prepended. The direction of the residual errors also differs. In both DR. INFO configurations, the predominant error was overtriage, which assigns a higher priority than the gold standard and therefore shortens the assessment interval rather than extending it. Overtriage and undertriage are not clinically equivalent: overtriage carries a resource cost but does not delay care for the patients most likely to deteriorate, whereas undertriage delays assessment at the priorities where delay carries the greatest risk of harm. The two retrieval augmented configurations therefore differ from GPT-5.1 in both the magnitude and the clinical direction of their errors.

Although both DR. INFO configurations matched on the overall undertriage rate without the anchoring statement (11.5%), the addition of the Manchester Triage System reference text to the agentic system’s retrieval layer produced incremental but consistent further improvement across nearly every other safety relevant metric. Exact match accuracy was higher with MTS retrieval than without (44.9% vs 40.4% without the anchoring statement, 55.8% vs 53.8% with it). Orange priority undertriage was lower (18.8% vs 21.9% without the anchoring statement, 12.5% vs 15.6% with it). Severe undertriage events fell from 2 to 1 without the anchoring statement and from 2 to 0 with it. Physician adjudicated agreement was higher (90.0% vs 83.3%, Section 3.3). Across these three reasoning regimes, the standalone language model showed the highest MTS undertriage rate on these vignettes, the agentic retrieval augmented system over a curated clinical knowledge base showed substantially lower undertriage, and the addition of the framework reference text produced further consistent improvement across multiple metrics. DR. INFO overtriaged 48.1% (Baseline) and 43.6% (with MTS retrieval), compared with 5.1% for GPT-5.1.

### 4.3 Anchoring resistance in the DR. INFO family

Under the misleading GP referral, both DR. INFO configurations held 0 of 8 Red-priority prompts undertriaged, matching their performance without the referral. GPT-5.1 undertriaged 8 of 8 Red-priority prompts (from 6 of 8 without the referral). DR. INFO Baseline and DR. INFO with MTS retrieval performed identically on this endpoint, so the resistance sits with the DR. INFO family, not with MTS retrieval specifically. The anchoring statement reproduces a routine deployment condition, in which clinical presentations arrive alongside referral letters, prior diagnoses, triage notes, and other clinician-authored framing that a triage adjunct must weigh against the primary clinical data. This is consistent with the established literature on anchoring as a cognitive bias in human clinical decision making (Croskerry, 2013).

### 4.4 Effect of adding more clinical context

The second property is an appropriate change in the assigned MTS priority when objective clinical data become available. The discriminators of the Manchester Triage System are operationally defined and threshold based. Examples include an abnormal pulse, a systolic blood pressure below 90 mmHg, and an SpO_2_ below 95%. When vital signs, examination findings, and laboratory values are present in the input, a system that retrieves these discriminators can compare the measured values against the thresholds defined by the Manchester Triage System. When the same case was presented with and without the objective data block added, DR. INFO with MTS retrieval was the only system whose undertriage rate changed significantly, with reductions distributed across the priority strata in which undertriage occurred. DR. INFO Baseline moved in the same direction with a smaller and non-significant change. GPT-5.1 moved in the opposite direction. This corresponds to standard triage practice (Mackway-Jones et al., 2014), in which a nurse assigns an initial level on partial information at first contact and revises it as vital signs, examination findings, and laboratory results accumulate. Reassessment in the waiting room is part of standard workflow across all five level scales (Manchester Triage System, ESI, CTAS) because a Yellow patient can become Orange or Red as the trajectory worsens. A triage adjunct whose assignment does not change in response to incremental clinical information cannot fulfil this requirement, and is therefore not suitable for a workflow in which repeated reassessment is the standard of care. The magnitude of the within-system change was: DR. INFO with MTS retrieval reduced its undertriage rate by 15.4 percentage points (19.2% to 3.8%, *p* = 0.005), DR. INFO Baseline reduced its undertriage rate by 2.6 percentage points (12.8% to 10.3%, *p* = 0.803), and GPT-5.1 moved in the opposite direction with a 6.4 percentage point increase (41.0% to 47.4%, *p* = 0.519). We tested the difference-in-differences directly by fitting a logistic regression with a system by variant interaction on the undertriage outcome. The interaction was significant for DR. INFO with MTS retrieval versus GPT-5.1 (*p* = 0.005), indicating that the two systems responded to the objective data block in significantly different directions, and did not reach significance for DR. INFO with MTS retrieval versus DR. INFO Baseline (*p* = 0.064), indicating that we cannot claim a significantly larger reduction for the MTS retrieval configuration on this sample. The pattern across the three within-system comparisons is that MTS retrieval is the only configuration for which the reduction in undertriage between the variant without objective data and the variant with it reached significance at the pre specified threshold. The benefit of objective clinical data was attenuated in all three systems when the anchoring statement was prepended, indicating that an authoritative but incorrect framing can partially override later corrective evidence, even in the system that otherwise updates on objective data correctly.

### 4.5 Two requirements for a safe AI triage adjunct

Two requirements emerge from these results for an AI triage adjunct that is suitable for clinical use:

1. A clinically conservative assignment when the case is first presented. At first contact, before vital signs, examination findings, and laboratory results have been gathered, the system has only the clinical history to work from. In this setting, the priorities at greatest risk of harm if delayed are Red and Orange presentations, and a safe adjunct must not assign them a lower priority than the gold standard.
2. An appropriate change in the assigned MTS priority when additional clinical information becomes available, with the change in the direction indicated by the Manchester Triage System discriminators. The triage assignment in clinical practice is not a one time decision but is revised as objective data accumulate. A system whose assignment does not change when those data are added to the input cannot reproduce this revision step.

In this benchmark, both DR. INFO configurations assigned zero of eight Red-priority prompts (derived from 2 Red cases) to a lower priority across every condition tested, whereas GPT-5.1 did so in the majority of Red-priority prompts. At Orange, both DR. INFO configurations undertriaged around 20% of prompts (14/64 Baseline, 12/64 with MTS retrieval) while GPT-5.1 undertriaged 47/64. DR. INFO therefore met requirement (1) at Red across all conditions and reduced but did not eliminate undertriage at Orange. On requirement (2), DR. INFO with MTS retrieval showed the largest reduction in undertriage when objective clinical data were added (19.2% to 3.8%). DR. INFO Baseline showed a smaller reduction in the same direction (12.8% to 10.3%). GPT-5.1 moved in the opposite direction (41.0% to 47.4%).

### 4.6 Case-level analysis of the general purpose model’s failure mode

Two cases illustrate the contrast in how the three systems handled the same evolving clinical evidence. In an acute asthma exacerbation, GPT-5.1 assigned Yellow despite SpO_2_ of 93–94%, peak flow at 62% of personal best, and a rising pCO_2_. Reassuring narrative phrases such as “still speaking in full sentences” and “not in obvious respiratory failure” appeared to drive the assignment, despite a clinical trajectory that the Red Zone criteria of the National Heart, Lung, and Blood Institute (NHLBI) classify as warranting immediate hospital referral. DR. INFO with MTS retrieval assigned Orange in three of the four permutations of this case and Red in one, and the response text named the Low SpO_2_ discriminator (defined in the Manchester Triage System Asthma flowchart as a saturation below 95% on air) and the “no improvement with own asthma medications” discriminator as positive, while explicitly noting that the peak flow of 62% of personal best did not meet the Low PEFR threshold (below 50%) (Mackway-Jones et al., 2014); DR. INFO Baseline reproduced the same Orange assignment in three of the four permutations and a Red overtriage in one. In a diabetic ketoacidosis case, GPT-5.1 identified an “early or mild DKA” in its response text but recommended an outpatient disposition, drawing on individual borderline values rather than on the trajectory of the laboratory and vital sign data taken together. DR. INFO with MTS retrieval assigned Orange in three of the four permutations and Red in one, and the response text named the Hyperglycaemia with ketosis discriminator from the Manchester Triage System Diabetes flowchart as positive (Mackway-Jones et al., 2014); DR. INFO Baseline reproduced the same pattern. Both GPT-5.1 cases point in the same direction as the failure to update on evolving clinical information reported by Ramaswamy et al. for ChatGPT on consumer queries (Ramaswamy et al., 2026), and are consistent with McCoy et al. (2025), who reported similar limitations in language models on clinical reasoning tasks (McCoy et al., 2025). The same mechanism is consistent with the direction of the objective data effect for GPT-5.1 in Section 3.2: when the objective data block was added to the same emergency cases, the model’s overall undertriage rate rose rather than fell, and the increase was concentrated at the highest acuity priorities, consistent with the model anchoring on reassuring individual values rather than synthesising the clinical trajectory.

### 4.7 Relation to concurrent work on clinical retrieval augmented systems

A concurrent evaluation in Nature Medicine compared three frontier general purpose language models against two commercial clinical retrieval augmented tools (OpenEvidence and UpToDate Expert AI) on a panel of clinical knowledge, expert alignment, and real world physician query benchmarks, and reported that the frontier models outperformed the commercial clinical tools across these tasks (Vishwanath et al., 2026). Those benchmarks evaluate broad clinical knowledge synthesis, in which the retrieved content is published literature that the language model must integrate into a free text clinical answer. The MTS triage task evaluated here is structurally different. Triage assignment against a published framework rewards faithful execution of an operationally defined decision procedure, and the retrieved content in DR. INFO with MTS retrieval is the procedure itself, with explicit discriminators and quantitative thresholds against which measured clinical values can be checked. The two findings therefore apply to different retrieval regimes. Vishwanath et al. themselves note that deeply subspecialised medical tasks may favour more sophisticated domain specific adaptation, and the MTS triage task evaluated here is one such setting, in which a curated clinical knowledge base alone closed most of the safety gap from a standalone frontier model and the addition of the framework reference text produced further consistent improvement across multiple safety relevant metrics.

## 5 Limitations

This study has limitations. First, language model outputs are stochastic, and the analysis is based on a single run per prompt per system per condition. Small count outcomes such as severe undertriage are therefore more sensitive to run to run variation than the overall undertriage contrast. Second, the sample of 39 cases yields only eight permutations at each of the Red and Blue priorities, so the confidence intervals at these levels are wide and the corresponding results are descriptive rather than inferential. Third, the pre-specified gender contrast was not significant in any system under either condition, but the gender comparison was limited to the two gender labels (man, woman) used in the original Ramaswamy et al. case set. Fourth, DR. INFO is an agentic retrieval-augmented system and GPT-5.1 was evaluated as a standalone model considered analogous to ChatGPT. The two systems therefore differ in more than a single component, and the cross-system contrast cannot isolate the contribution of any specific element. The within-system contrast between DR. INFO Baseline and DR. INFO with MTS retrieval is unaffected by this consideration, because both configurations share the same underlying system apart from the MTS retrieval layer. Fifth, the MTS-Bench dataset was constructed by the team that developed DR. INFO, a configuration that Vishwanath et al. have noted may systematically favour the systems being evaluated (Vishwanath et al., 2026). The within-system contrast and the independent physician adjudication described in Section 3.3 are intended to mitigate this concern in the present study; independent replication on a benchmark constructed outside any commercial entity remains the cleanest external test. Sixth, the physician adjudication was conducted by a single physician on the high risk subset; multi-rater adjudication and extension to the full case set are planned for follow up work. Seventh, the 156 permutations per system per condition are not independent observations, since they arise as 4 permutations of 39 underlying cases. The primary Fisher’s exact test therefore treats correlated permutations as independent, which is anti conservative for the between-system contrasts. Sensitivity analyses at the case level (Fisher’s exact on majority undertriaged cases; GEE logistic regression clustered by case) produced the same qualitative direction and significant differences, but a fully case-clustered primary analysis with a larger case pool is a natural next step. The within-system objective-data (E versus F) and gender comparisons are also paired at the case level, and Fisher’s exact test treats those observations as independent; a paired test at the case level would be more powerful for those contrasts. Eighth, the gold-standard priorities were derived by our annotation pipeline against the Manchester Triage Group reference text (Mackway-Jones et al., 2014), and the DR. INFO with MTS retrieval configuration retrieves passages from the same text at inference time. The vignettes themselves are not present in the reference text, and the retrieval configuration was not exposed to the gold-standard priorities produced by the annotation pipeline; both processes independently apply the framework’s discriminators to each vignette. The MTS retrieval configuration’s agreement with the gold standard therefore reflects consistency with the Manchester Triage System framework, not agreement with an independent clinical outcome. Comparisons of DR. INFO Baseline against GPT-5.1 are not affected. Ninth, this benchmark measures MTS priority agreement on physician-authored vignettes; it does not measure patient outcomes or deployment safety, and aggregate benchmark performance may obscure error patterns with clinical implications.

## 6 Conclusion

This benchmark evaluated three reasoning regimes for an AI triage adjunct against the Manchester Triage System on 312 physician style vignette prompts per system. OpenAI GPT-5.1, accessed via API and used as a standalone triage adjunct, showed a similar undertriage pattern to that previously reported for ChatGPT (Ramaswamy et al., 2026), with systematic undertriage of the most time critical presentations that was amplified when a misleading GP referral statement was prepended. Replacing the standalone language model with an agentic retrieval augmented system over a curated clinical knowledge base (DR. INFO Baseline) substantially reduced the safety relevant errors, with 0 of 8 Red-priority prompts undertriaged in every condition tested. Adding retrieval of the Manchester Triage System textbook (DR. INFO with MTS retrieval) produced the largest reduction in undertriage when objective clinical data were added, in the direction indicated by the Manchester Triage System discriminators. Neither DR. INFO Baseline nor GPT-5.1 showed a comparable change.

Of the three configurations evaluated in this benchmark, only DR. INFO with MTS retrieval met both requirements identified in Section 4.5: a clinically conservative assignment at first contact, and an appropriate change in the assigned priority when objective clinical data become available. The confidence intervals at the Red priority remain wide because only eight prompts, derived from two Red cases, contribute at that level. MTS-Bench provides a standardised framework for evaluating language model based triage safety against an established nurse led emergency triage standard. Prospective evaluation in working emergency department and primary care settings, with clinicians using the system in real workflow, is the necessary next step.

## Data Availability

MTS-Bench is available from the corresponding author on reasonable request. The clinical scenarios from which MTS-Bench was adapted are published by Ramaswamy et al. (2026, Nature Medicine, doi:10.1038/s41591-026-04297-7). The Manchester Triage Group reference text used as the gold standard is published by Mackway-Jones et al. (2014, John Wiley & Sons/BMJ Books, ISBN 978-1-118-29906-7).

## Acknowledgements

We gratefully acknowledge the technical support provided by Sonu Kumar and Kritika Singh.

## Data availability

MTS-Bench is available from the corresponding author on reasonable request.

